# Efficacy of remdesivir versus placebo for the treatment of COVID-19: A protocol for systematic review and meta-analysis of randomized controlled trials

**DOI:** 10.1101/2020.04.09.20059196

**Authors:** Desye Gebrie, Desalegn Getnet, Tsegahun Manyazewal

## Abstract

**Background:** In spite of the global containment on prevention efforts, the spread of coronavirus disease 2019 (COVID-19) is continuing to rise, with 1.1 million confirmed cases and 60,124 deaths recorded worldwide since 04 April 2020. The outbreak has a significant threat to international health and economy. At present, there is no approved vaccine or treatment for the disease, while efforts are underway. Remdesivir, a nucleotide-analogue antiviral drug developed for Ebola, is determined to prevent and stop infections with COVID-19, while results are yet controversial. Here, we aim to conduct a systematic review and meta-analysis of randomized controlled trials to compare the effectiveness of remdesivir and placebo in patients with COVID-19.

**Method and analysis:** We will search MEDLINE-PubMed, Embase, Cochrane Library, ClinicalTrials.gov, and Google scholar databases without restriction in year of publication. We will include randomized controlled trials that assessed the effectiveness of remdesivir versus placebo for patients confirmed with COVID-19. We will follow the Preferred Reporting Items for Systematic Review and Meta-Analysis (PRISMA 2015) guidelines for the design and reporting of the results. The primary endpoint will be time to clinical recovery. The secondary endpoints will be all cause mortality, discharged date, frequency of respiratory progression, and treatment-emergent adverse events. Two independent authors will perform study selection, data extraction, and methodology quality assessment. RevMan 5.3 software will be used for statistical analysis. Random/fixed effect model will be carried out to calculate mean differences for continuous outcomes and risk ratio for dichotomous outcomes between remdesivir and placebo.

**Ethics and dissemination:** This study does not require ethical approval, because no participant’s data will be involved in this systematic review and meta-analysis. The findings of this study will be published in reputable and peer-reviewed journal.

**Registration:** This review protocol is submitted in PROSPERO database for registration and we will include the registration number in the revised version of the manuscript.

**Strengths and limitations of this study:**

➣ This systematic review and meta-analysis will be derived from only randomized controlled trials which will increase the quality of evidences.
➣ This systematic review and meta-analysis will be derived from only randomized controlled trials which will reduce between study heterogeneity.
➣ Subgroup and sensitivity analysis will be carried out to identify possible reasons that may cause significant heterogeneity between studies.
➣ The use of Cochrane risk of bias tool to assess risk of bias for each included studies to extract and synthesize evidence based conclusions.
➣ One of the limitation of this study might be the restriction of trials published in English language.

## Introduction

Over the course of December 2019, the health authority of Wuhan City, Hubei province, China reported a cluster of pneumonia cases of unknown etiology [1]. The Chinese researcher rapidly isolated Sever Acute Respiratory Syndrome Coronavirus 2 (SARS-CoV-2) from a patient on 7 January 2020 and came out to genome sequencing of the SARS-CoV-2 [2]. On 9 January 2020, China’s communicable diseases control authority announced that 2019 novel coronavirus (2019-nCoV) had been detected as the causative agent for the epidemics [3]. On 11 February 2020 World Health Organization officially named the disease as coronavirus disease 2019 (COVID-19). COVID-19 is caused by a novel β-coronavirus which is named as SARS-CoV-2. SARS-CoV-2 shares 79% sequence identity with Sever Acute Respiratory Syndrome Coronavirus (SARS-CoV) and Middle East Respiratory Syndrome Coronavirus (MERS-CoV) which caused a major outbreak since 2002 and 2012 in China and Saud Arabia respectively [4-6].

In spite of the global containment on prevention efforts, the spread of COVID-19 is continuing to rise with 1.1 million confirmed cases and 60,124 deaths recorded worldwide since 04 April 2020. [7-8]. The outbreak of COVID-19 infection has a significant threat to international health and economy [9]. At present, there is no approved vaccine or treatment for COVID-19, so that identifying the drug treatment options as soon as possible is critical agenda to overcome the outbreak [10-11].

Despite the lack of approved drugs and vaccine for COVID-19, many scientists are endeavoring to find medicines specific to the virus and they have been looking into repurposing the already approved drugs. As of 29 March 2020, there has been 209 clinical trials registered in clinicaltrials.gov and estimated to be over 500 [12]. Currently, several drugs such as remdesivir, hydroxychloroquine, chloroquine, Ritonavir+Lopinavir, Arbidol and interferon are undergoing randomized controlled trials (RCTs) to test their efficacy and safety for the treatment of COVID-19 in many countries [13-18]. Among these investigating drugs remdesivir showed promising results [18-19]. Remdesivir is nucleotide analog prodrug and shows broad spectrum antiviral activity against many RNA viruses including SARS-CoV-2 [20-21]. Remdesivir has been reported as a treatment of COVID-19 in United States, China and Italy [13,15, 22]. while results are yet controversial [9]. To bridge this gap, here we aim to conduct a systematic review and meta-analysis of RCTs to compare the effectiveness of remdesivir and placebo in patients with COVID-19.

## Methods

### Study registration

The protocol for this systematic review and meta-analysis is submitted in PROSPERO database for registration and we will include the registration number in the revised version of the manuscript.

### Data sources and searches

We will search MEDLINE/PubMed (http://www.ncbi.nlm.nih.gov/pubmed/), Embase (http://www.embase.com/), The Cochrane Library (http://www.cochranelibrary.com/), ClinicaTtrials.gov (https://www.clinicaltrials.gov/), and google scholar (https://scholar.google.com/) databases for completed studies that reported the efficacy of remdesivir versus placebo for patients with COVID-19. We will include randomized controlled trials that assessed the effectiveness of remdesivir versus placebo for patients with COIVID-19 without restriction on year of publication, but published in English language. The Medical Subject Headings (MeSH) and keywords we will used in different combinations using balloon operators will be 2019 novel coronavirus, 2019-nCov, coronavirus disease 2019, COVID-19, SARS-cov-2, remdesivir, nucleotide-analogue, antiviral drug and randomized controlled trials. All potentially eligible studies will be considered for this review, irrespective of the primary outcomes. Manual searching will be performed to find out additional eligible trials from the reference lists of key articles.

**Table 1:**
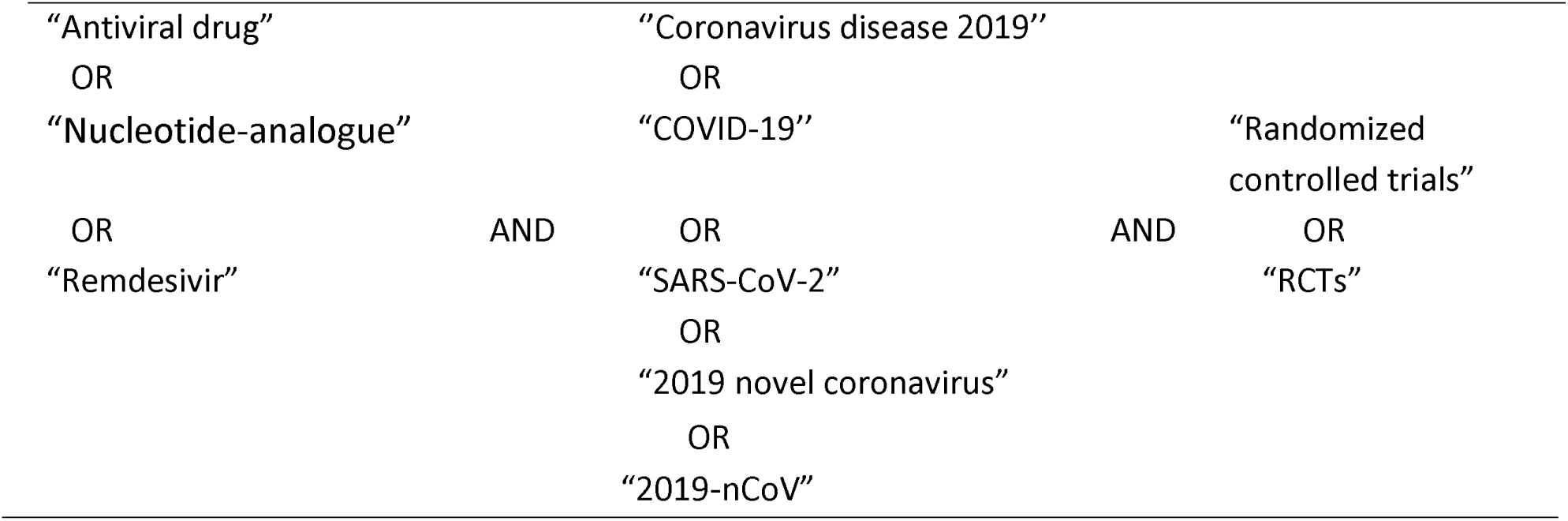
Search strategy for the MEDILINE-PubMed database

### Eligibility

Study eligibility criteria for this systematic review and meta-analysis will be in accordance with Participants, Intervention, Comparison, Outcomes and Study designs (PICOS) descriptions [23].

### Population

The population will be patients confirmed with COVID-19 and with or without other co-morbid conditions in all age groups.

### Intervention

The intervention/ experimental group will be any dose of remdesivir

### Comparator

The comparator group will be placebo/ standard of care

### Outcomes

The primary endpoints will be time to clinical recovery and proportion of participants relieved from clinical symptoms defined at the time (in hours) from initiation of the study treatment. The secondary endpoints will be all cause mortality, discharged date, frequency of respiratory progression, oxygen saturation and treatment-emergent adverse events in each groups.

### Study design

Only RCTs evaluating effectiveness of remdesivir versus placebo for patients with COVID-19 will be included.

### Study selection

The title and abstract of all searched studies will be examined by two independent review authors. From the title and abstract of all studies identified by the database search, those studies duplicated and not meet the eligibility criteria will be excluded. The full texts of the remaining studies will be further reviewed. Disagreements will be resolved by consensus and if persisted, we will be arbitrated through discussion with a third review author. We will follow the Preferred Reporting Items for Systematic Review and Meta-Analysis (PRISMA 2015) guidelines [24] for the design and reporting of the results.

**Figure 1:**
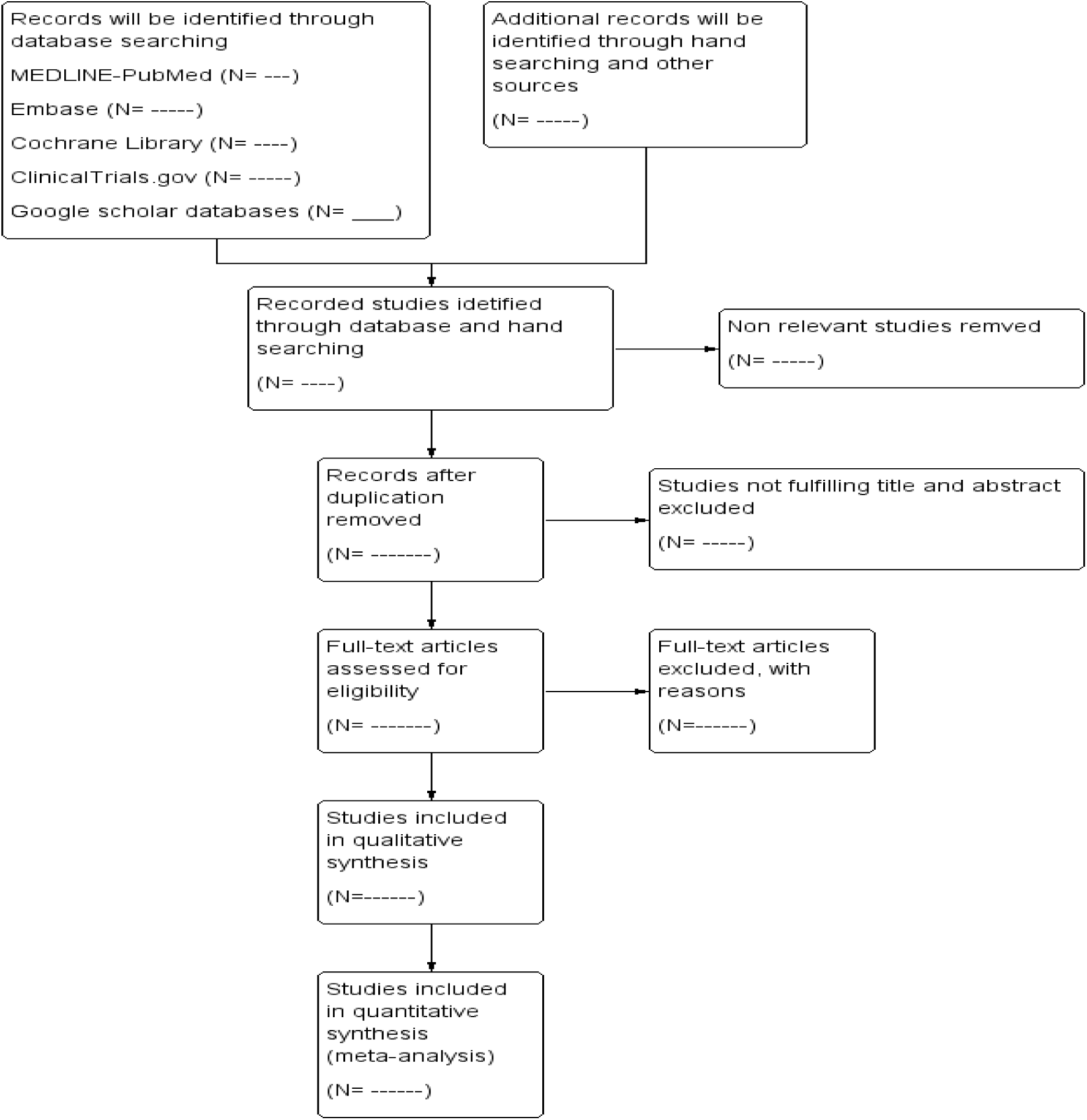
Preferred Reporting Items for Systematic Reviews and Meta-Analyses flow diagram of the study selection process and search results

### Data extraction

Two authors will independently extract data according to the pre-designed data extraction tool. The following data will be extracted from each included RCTs: first author, year of publication, funding information, setting, mean age of the participant, interventions, comparators, doses, number of participants randomized, duration of treatment, all primary, secondary and other outcome measurements. If any disagreement regarding the data extraction between the two review authors exist, the third author will be consulted and consensus will be made through discussion.

**Table 2:** characteristics of RCTs included in the systematic review and/or meta-analysis

| 1<sup>st</sup> author<br>(year) | Country | Age<br>(year) | No.<br>of<br>pts | Intervention | Comparator | Follow<br>-up<br>(days) | Outcomes | Results |  |
| --- | --- | --- | --- | --- | --- | --- | --- | --- | --- |
|  |  |  |  |  |  |  |  | Remdesivir | Placebo |
|  |  |  |  | Remdesivir<br>(n=) | Placebo (n=) |  | Time to clinical<br>recovery<br>No. of patients<br>relieved from<br>symptoms<br>Frequency of<br>respiratory<br>progression<br>Oxygen<br>saturation<br>Adverse events<br>Death events |  |  |

### Assessment of risk of bias

The Cochrane risk of bias tool [25] will be used to assess the risk of bias for each included study. The risk of bias of each trial will be judged by two independent review authors as “Low”, “Unclear”, or “High” based on the critical domains, including random sequence generation, allocation concealment, blinding, incomplete outcome data, selective reporting and other source of biases. Disagreements will be resolved by discussion among all authors. If the disagreements cannot be resolved through discussion, an arbiter will make the final decision.

### Statistical analysis

Meta-analysis will be carried out using the computer software packages RevMan 5.3 [26]. Continuous outcome data will be reported using a mean difference (MD) and a 95% confidence interval (CI). Binary outcome data will be summarized using risk ratio (RR) and 95% CI. Mantel-Haenszel method [27] will be used to pool effect estimates of dichotomous outcomes and inverse variance for continuous outcomes. Cochrane Q test [28] will be used to assess heterogeneity between studies, and I^2^ testing [29] will be done to quantify heterogeneity between studies, with values > 50% representing moderate-to-high heterogeneity. If heterogeneity between study is acceptable, a fixed-effect model will be used to pool the data. On the other hand, if unacceptable heterogeneity detected or if the number of studies are small, a random-effect model will be used to pool the data [30]. Subgroup analysis will be carried out to identify possible reasons that may cause significant heterogeneity between studies. If we get acceptable heterogeneity after the subgroup analysis, we will perform meta-analysis. Otherwise, we will do a narrative description. Sensitivity analysis will be conducted to see the robustness of pooled data by removing low quality studies. Statistical analysis with a p-value < 0.05 will be considered statistically significant.

### Addressing missing data

When individual participant’s data are initially unavailable, we will review the original source, and/or published trial reports and we will contact the authors to obtain clarification for these data.

### Reporting bias

We will conduct funnel plot and Egger test to check any possible reporting bias if a sufficient number of included studies (at least 10 trials) are available in this study [31].

## Data Availability

all important data are available within the manuscript.

## Ethics and dissemination

This study does not require ethical approval, because no participant’s data will be involved in this systematic review and meta-analysis. The findings of this study will be published in reputable and peer-reviewed journal.

## Abbreviations

2019-nCoV: 2019 novel Coronavirus
COVID-19: Coronavirus Disease-2019
SARS: Sever Acute Respiratory Syndrome
RCTs: Randomized Controlled Trials
SARS-CoV-2: Sever Acute Respiratory Syndrome Coronavirus-2
MERS: Middle East Respiratory Syndrome

## Declarations

### Authors’ contributions

DG (first author) conceived the study, developed the study criteria, searched the literature, wrote the protocol and drafting the manuscript. DG (second author) conducted the preliminary search and TM revised the manuscript. All authors have read and approved the manuscript.

## Acknowledgement

The authors would like to acknowledge Center for Innovative Drug Development and Therapeutic Trials for Africa (CDT-Africa), College of Health Sciences, Addis Ababa University which is funding this study.

